# Functional Correlates of Cognitive Performance and Working Memory in Temporal Lobe Epilepsy: Insights from Task-based and Resting-state fMRI

**DOI:** 10.1101/2022.08.02.22278343

**Authors:** Alfonso Fajardo-Valdez, Vicente Camacho-Téllez, Raúl Rodríguez-Cruces, María Luisa García-Gomar, Erick H. Pasaye-Alcaraz, Luis Concha

## Abstract

Temporal lobe epilepsy (TLE) is a common form of medically intractable partial epilepsy. Although seizures originate in mesial temporal structures, there are widespread abnormalities of gray and white matter beyond the temporal lobes that negatively impact on functional networks and cognition. Previous studies have focused either on the global impact on functional networks, or on the functional correlates of specific cognitive abilities. Here, we use a step-wise approach to evaluate the link between whole-brain functional connectivity (FC) anomalies to overall cognitive performance, and how such abnormal connectivity alters the fronto-parietal brain regions involved in working memory (WMem), a cognitive disability often reported by TLE patients. We evaluated 40 TLE patients and 36 healthy subjects through extensive cognitive testing, resting-state functional magnetic resonance imaging (rs-fMRI), and task-based fMRI using Sternberg’s task to evaluate WMem. As a group, TLE patients displayed cognitive abnormalities across different domains, although considerable within-group variability was identified. TLE patients showed disruptions of functional networks between and within the default mode network (DMN) and task-positive networks (TPN) resulting in associations with cognitive performance. Furthermore, during the WMem task, TLE patients showed abnormal activity of fronto-parietal regions that was associated with other forms of memory, and alterations of seed-based connectivity analyses. Our results show that different degrees of abnormal functional brain activity and connectivity are related to the severity of cognitive disabilities across cognitive spheres. Differential activity between patients and healthy subjects suggest potential compensatory mechanisms to preserve adequate cognitive performance.

## 1. Introduction

Temporal lobe epilepsy (TLE) is the most common type of focal epilepsy (Téllez-Zenteno & Hernández-Ronquillo, 2012). TLE seizures are often refractory to antiepileptic drugs (AEDs), and due to this, TLE usually entails large detrimental effects on the quality of life of patients, affecting their physical and mental health, with a wide spectrum of cognitive deficits (Giovagnoli & Avanzini, 2000; Khalife et al., 2022). Long-term memory deficits are one of the most frequent concerns in these patients (Fisher et al., 2000; Hoppe et al., 2007), with self-reported memory being an important predictor of quality of life that correlates with objective performance on memory tests (Giovagnoli & Avanzini, 2000). Notably, cognitive deficits are already present in newly-diagnosed epilepsy in children (Khalife et al., 2022), with potential of further deterioration due to long-term pharmacological treatment (Eddy et al., 2011). A large body of research has reported that TLE patients typically undergo damage in their ability to recall, often displaying important short- and long-term memory deficits (Zeman et al., 2012). It is well known that the hippocampus and medial temporal lobes play critical roles in declarative memories (Milner, 1972; Scoville & Milner, 1957). Indeed, patients that exhibit neuronal loss, volume reduction, and gliosis in mesial temporal structures (mesiotemporal sclerosis, MTS) also display the most severe deficits in short- and long-term memory (Delaney et al., 1980; Zeman et al., 2012). Notably, structural and functional abnormalities are not restricted to the temporal lobe, with a large body of evidence showing alterations widespread throughout the brain, comprising neocortical thinning, atrophy of subcortical structures, and abnormalities of white matter (Deleo et al., 2018; Hatton et al., 2020; Whelan et al., 2018). Such widespread alterations are linked to deficits in cognitive domains not necessarily or exclusively linked to temporal lobe function, such as executive functions, attention, and working memory (Agah et al., 2017; Dabbs et al., 2009; Hudson et al., 2014; Rodríguez-Cruces et al., 2020; Stretton et al., 2013; Zamarian et al., 2011).

Resting-state functional connectivity (RSFC) has been used to show abnormalities of the default mode network (DMN), attention, and reward/emotion networks that may partially explain cognitive deficits across domains (Caciagli et al., 2014; Rodriguez-Cruces et al., 2022). Cognitive abilities of healthy participants are correlated with the adequate interplay of the DMN and task-positive networks (TPN) (Konishi et al., 2015; Smallwood et al., 2012). Nonetheless, it is unknown how the altered networks of TLE patients modulate the interaction between large-scale functional connectivity and specific cognitive abilities.

The concept of working memory (WMem) was first described by Miller in 1960 and refers to a type of short-term memory which has a crucial cognitive function that supports ongoing and upcoming behaviors, allowing storage of information across delay periods (Mansouri et al., 2015). A basic feature of WMem is the maintenance of information in the short term in absence of any sensory input (Eriksson et al., 2015). This means that once the information is presented, the individual is able to retain and manipulate it without the stimulus remaining present. WMem is a basic component of complex cognitive functions such as learning and reasoning (Baddeley, 2012). Crucially, WMem does not appear to involve temporal lobe structures, but rather maintenance of information in WMem is carried out within a fronto-parietal network (Eriksson et al., 2015). WMem appears to be further impaired in patients with left TLE with hippocampal sclerosis, early onset of the disorder, and high frequency of seizures (Zhao et al., 2014). Functional Magnetic Resonance Imaging (fMRI) has shown bilateral activation of the frontal and parietal lobes using paradigms to evaluate WMem (e.g., N-back) (Owen et al., 2005) and such activity is impaired in TLE patients (Stretton et al., 2013; Vlooswijk et al., 2011). In healthy subjects, the hippocampus is normally bilaterally deactivated during WMem tasks, and the level of such deactivation is dependent on memory load. Interestingly, this pattern is abnormal in patients with TLE (Stretton et al., 2012). While group studies have shown that TLE patients have WMem deficits (Hermann et al., 2007; Stretton et al., 2012; Tudesco et al., 2010; Vlooswijk et al., 2011), the impact of WMem network activity on overall cognitive performance of TLE patients is unknown.

In this work, we explored the association between brain activity and cognitive performance in TLE patients using two complementary approaches. First, we explore the link between cognitive abilities across domains with the activity of resting-state functional networks. Next, we use task-based fMRI to specifically investigate WMem abilities and fronto-parietal network activity. Finally, we investigate the interplay between WMem networks resulting from task-fMRI and those derived from resting-state fMRI, and how both impact on overall cognitive abilities.

## 2. Materials and Methods

### 2.1. Participants

We recruited a sample of 40 patients with TLE, previously reported in (Rodríguez-Cruces et al., 2020) (age 31 ± 11.6 years old, 14 women). Patients were diagnosed and followed at out-patient epilepsy clinics at Hospital General de México in Mexico City, and at Hospital Central “Dr. Ignacio Morones Prieto” in San Luis Potosí, México. The referring neurologists diagnosed the patients according to ILAE criteria, relying on clinical information, surface EEG, and conventional neuroimaging. Based on electroclinical and neuroimaging data, the laterality of the epileptogenic focus was determined, resulting in 20 left-TLE, 16 right-TLE, and 4 bilateral TLE patients. The control group consisted of 36 healthy subjects matched for age and education (age 33 ± 12 years old; 17 women) without any history of neurologic or psychiatric disease. All participants were right-handed. Therefore, as mentioned The study was approved by the Ethics Committee of the Institute of Neurobiology and written informed consent was obtained from all subjects in accordance with the Council for International Organizations of Medical Sciences (CIOMS) International Ethical Guidelines for Health-Related Research Involving Human Subjects.

### 2.2. Neuropsychological assessment

Participants underwent extensive cognitive assessment at Hospital General de México and Centro Estatal de Salud Mental, Querétaro, México, which was conducted by experienced neuropsychologists. Two scales were used: Wechsler’s Adult Intelligence Scale (WAIS-IV) and Weschsler’s Memory Scale (WMS-IV). Scores of these tests were reported as percentiles adjusted for age and education level based on the Mexican population. Derived from these two scales, we used nine indices as measurements of performance on specific cognitive domains: verbal comprehension (VCI), perceptual reasoning (PR), processing speed (PSI), working memory (WMI), visual memory (VMI), visual working memory (VWMI), immediate memory (IMI), delayed memory (DMI), and auditory memory (AMI).

### 2.3. Magnetic Resonance Imaging

We acquired resting-state (RS) and task-fMRI sequences. Studies were performed on a 3 T Philips Achieva TX scanner (Best, The Netherlands) at the National Laboratory for MRI, located at Instituto de Neurobiología, UNAM Campus Juriquilla, Querétaro, Mexico. For the acquisition of RS-fMRI, participants were instructed to close their eyes, stay still and relaxed, and to stay awake during the scanning session. Gradient-echo echo-planar T2*-weighted images (TR/TE = 2000/30 ms) were acquired covering the whole brain with 34 slices, with a voxel resolution of 2×2×3 mm^3^ (200 volumes; 6 minutes 40 seconds). Similarly, task-fMRI (190 volumes; 6 minutes 20 seconds) was acquired with the same scan parameters while subjects performed the Sternberg memory task (described below). For registration purposes, we also acquired high-resolution T1-weighted images with 1×1×1 mm^3^ voxel size using a 3D magnetization-prepared gradient echo sequence (TR/TE=8.1/3.7 ms; flip angle=8°). Additionally, other structural and DWI sequences were acquired and described elsewhere (Rodríguez-Cruces et al., 2018, 2020). We carried out quality assessment on fMRI images, and exclusion criteria for subjects was independent for each modality. Therefore, the number of included subjects differ slightly between RS-fMRI and task-fMRI analyses and are reported on each section (these subsamples did not differ on sex, age and education levels).

### 2.4. Sternberg’s memory task

We used a modified version of Sternberg’s memory task (Black et al., 2010) which was introduced to participants prior to the imaging session, making sure they understood and complied with the task. Participants lied on the scanner and viewed a series of slides through a mirror affixed to the MRI head coil. Each trial consisted of three slides (Figure 1), which were presented using E-prime version 2 (Psychology software tools, Sharpsburg, PA): The first slide showed a list of either 2, 4, or 6 digits which participants were instructed to memorize (encoding phase, 4 s duration); the second slide showed only three dots that indicated that the previous list of digits must be retained in memory (retention phase, 10 s); the last slide showed a single digit between question marks, and the subject was asked to respond whether that specific digit was included in the list of digits seen in the first slide (retrieval phase, 4 s). A total of 12 trials with different memory loads and digits in each list were evaluated per participant, with an inter-trial period of 18 seconds. Participants provided their response (yes/no) through MRI-compatible triggers (Nordic NeuroLab, Bergen, Norway) by pressing a button with either index finger; the side indicating yes or no was counterbalanced between participants. Participants were instructed to provide their responses as quickly as possible. The number of correct responses and the reaction times were analyzed as measures of performance in the task, through mixed 2×3 analyses of variance (ANOVA) considering group (TLE or Control) as the between-subjects factor and memory load (2, 4, or 6 digits) as the within-subjects factor. statistical significance was considered at a *p <* 0.05.

**Figure 1:**
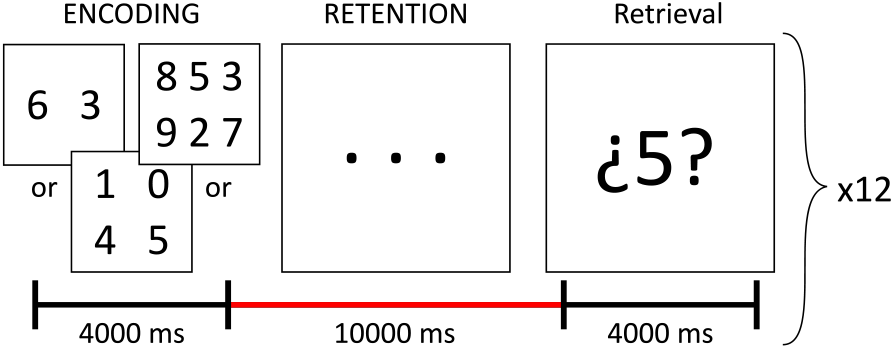
Sternberg memory task for evaluation of WMem.

### 2.5. Resting-State fMRI preprocessing

RS-fMRI datasets were preprocessed with the Configurable Pipeline for the Analysis of Connectomes (C-PAC v.1.1.8) (Craddock et al., 2013), set with the default parameters with minimal adjustments based on (Power et al., 2012; Satterthwaite et al., 2013). Preprocessing pipeline included the following procedures: slice-timing correction for an interleaved acquisition, motion correction, skull and non-brain tissue removal, alignment of functional timeseries to the anatomical T1-weighted volume, nuisance artifact removal (nuisance regression), spatial normalization and spatial smoothing. The matrix of nuisance regressors included linear and quadratic trends, white matter (WM) mean intensity, cerebrospinal fluid (CSF) mean intensity, the top 6 principal components derived from WM and CSF signal (aCompcor), motion parameters and their derivatives (Friston et al., 1996), and 0.01-0.08 Hz bandpass filtering. Additionally, framewise displacement (FD) was calculated from rigid motion parameters, and timepoints whose FD value was greater than 0.25 were marked as spikes and added to the nuisance regressors matrix (volume censoring) to prevent spurious results in posterior analyses. Following nuisance regression, residuals were spatially normalized and warped to the Montreal Neurological Institute 152 atlas (MNI152, voxel resolution of 2×2×2 mm^3^), and data was spatially smoothed through the application of a gaussian kernel of 6 mm FWHM. After visual quality assessment of preprocessed data, datasets with excessive head motion, defined as remaining with less than 120 non censored timepoints (i.e., less than 4 minutes of usable signal) were discarded from the study. As a result, RS-fMRI data from 45 subjects remained usable, from which 25 were controls (18 women), and 20 were patients (14 women,12 left-TLE, 8 right-TLE). Due to the large exclusion of TLE datasets, and to increase the sensitivity of analyses, we formed a homogeneous TLE group by flipping the brains (inverting the *x*-axis coordinates of images) of right TLE patients. Therefore, results of RS-fMRI analyses are referenced to the location of epileptogenic focus (ipsilateral/contralateral) rather than to the hemispheric (left/right) location of brain regions.

### 2.6. Definition of DMN and TPN regions of interest

Regions of interest (ROIs) of DMN and TPN were obtained from neurosynth.org (Yarkoni et al., 2011). Briefly, we selected the MNI152 seed voxel coordinates, *x* = *−*8, *y* = *−*56, *z* = 26, corresponding to the posterior cingulate cortex (PCC) (Andrews-Hanna et al., 2010) Seed coordinates were then used in the neurosynth’s locations tool to generate a seed-based Pearson coefficient correlation whole-brain map based on a sample of 1000 RS-fMRI datasets (see https://neurosynth.org/locations/ for more details). Next, said map was thresholded, keeping only *r* values lower than -0.25 or greater than 0.25, and as a result we obtained 21 clusters, 8 corresponding to DMN regions and the remaining 13 corresponding to task-positive regions (DMN anticorrelations). These clusters were used as a template to generate 4 mm radius spherical ROIs centered at each cluster’s local maxima (see Table 1).

**Table 1:** Regions of interest. Spatial coordinates (*x, y, z*) of local maxima in MNI space (mm).

| Region of interest (ROI) | Abbreviation | $x$ | $y$ | $z$ |
| --- | --- | --- | --- | --- |
| Contralateral Hippocampus | C.HIPP | 26 | -16 | -16 |
| Ipsilateral Hippocampus | I.HIPP | -24 | -18 | -16 |
| Contralateral Middle Temporal Gyrus | C.MTG | 62 | -4 | -16 |
| Ipsilateral Middle Temporal Gyrus | I.MTG | -62 | -8 | -14 |
| Contralateral Posterior Parietal Lobe | C.PPL | 52 | -60 | 32 |
| Ipsilateral Posterior Parietal Lobe | I.PPL | -50 | -66 | 34 |
| Posterior Cingulate Cortex | PCC | -8 | -56 | 26 |
| Ventromedial Prefrontal Cortex | VmPFC | 0 | 54 | -6 |
| Ipsilateral Middle Temporal Gyrus (Temporal-Occipital part) | I.MTG-to | -56 | -62 | 0 |
| Contralateral Middle Temporal Gyrus (Temporal-Occipital part) | C.MTG-to | 60 | -54 | -4 |
| Ipsilateral Precentral Gyrus | I.PrG | -28 | -6 | 52 |
| Ipsilateral Parieto-Occipital Sulcus | I.POS | -18 | -70 | 50 |
| Contralateral Parieto-Occipital Sulcus | C.POS | 18 | -70 | 54 |
| Paracingulate Gyrus | PCG | 4 | 12 | 52 |
| Ipsilateral Insula | I.INS | -32 | 16 | 8 |
| Contralateral Insula | C.INS | 34 | 18 | 6 |
| Ipsilateral Frontal Pole | I.FP | -38 | 42 | 34 |
| Contralateral Frontal Pole | C.FP | 38 | 46 | 30 |
| Contralateral Precuneus | C.PrC | 14 | -34 | 44 |
| Ipsilateral Supramarginal Gyrus | I.SMG | -62 | -36 | 38 |
| Contralateral Supramarginal Gyrus | C.SMG | 62 | -32 | 42 |

### 2.7. Functional connectivity and network analysis

Following preprocessing, RS-fMRI data from 46 subjects remained usable, from which 25 were controls (18 women), and 21 were patients (14 women,13 left-TLE, 8 right-TLE). Due to the large exclusion of TLE datasets, and to increase the sensitivity of analyses, we formed a homogeneous TLE group by flipping the brains (inverting the x-axis coordinates of images) of right TLE patients. Therefore, results of RS-fMRI analyses are referenced to the location of epileptogenic focus (ipsilateral/contralateral) rather than to the hemispheric (left/right) location of brain regions. The ROIs mean timeseries were extracted from each preprocessed RS-fMRI dataset, and we obtained 21×21 FC matrices through calculation of Pearson’s correlation coefficients between timeseries of each pair of ROIs. We then carried out statistical comparisons of FC between patients and controls, through node-based and network-based approaches. For the node-based approach, matrices were linearized and FC from each pair of nodes was compared between groups, yielding computation of multiple two-tailed t-tests for independent samples, followed by adjustment of *p* values with false discovery rate (FDR). A network-based approach was conducted using the Network-Based statistic (NBS) toolbox (Zalesky et al., 2010) on MATLAB v.2018a. Similarly to the cluster correction in voxelwise analyses, NBS focuses on the computation of cluster statistics rather than on independent node statistics, and thus, it yields a higher sensitivity and statistical power at the cost of reducing the specificity of statistical comparisons. To conduct NBS analysis, first, an initial cluster threshold value is chosen to identify significant connections between pairs of nodes. Next, connections that surpassed such threshold are used to reveal the distinct components of the network. Lastly, statistical comparisons between samples are carried out, and a *p* value is assigned to the components through a permutation test. In this study we tested between-groups hypotheses bidirectionally (Controls*>*Patients; Controls*<*Patients) with an initial threshold set at t-test *>*2.1 and subsequent permutations test set at *n* = 5000. In both the network-based and the node-based approaches, results were deemed significant at a *p <* 0.05.

### 2.8. Association between resting-state functional connectivity and neuropsychological scores

We aimed to examine whether FC of DMN-TPN correlated with performance on the different cognitive domains of each group. To achieve this, we fitted analyses of covariance (ANCOVA) models setting each cognitive domain as the predicted variable and each node-node FC value, group and [FC × group] interaction as the predictors of the model. ANCOVA models can be described with by the following linear model equation: cognitive score = *β*_0_ +*FC*_*ij*_ +*Group*+*FC*_*ij*_ × *Group*+*error*. We focused on testing the significance of the interaction term of the models to find specific group associations. Since we built an independent model for each functional connection, we controlled for false positives by calculating an adjusted *p* value threshold using Bonferroni’s method (*α/n*, with *α* = 0.05 and *n* = 9 cognitive indexes, resulting in *p*_*corr*_ *<* 0.0055.

### 2.9. General linear model of fMRI during Sternberg’s memory task

The analyzed sub-sample consisted of 66 subjects, from which 33 were controls (26 women) and 33 were TLE patients (25 women,18 left-TLE, 11 right-TLE, 4 Bi-TLE). Images were preprocessed with fMRIprep v.1.5.5 (Esteban et al., 2019). Preprocessing pipeline for task-fMRI sequences was similar to the RS-fMRI pipeline except that we applied high-pass filtering instead of bandpass filtering, and volume censoring and nuisance regression steps were not carried out. To evaluate the Wmem network we used the FEAT tool in FSL (version 6.00, FSL tools, FMRIB, Oxford) (Smith et al., 2004), and carried out a general linear model (GLM) for a block design on each subject’s image series. First-level analyses were carried out by modeling the BOLD signal time series as a linear combination of six main regressors: one regressor for each of the three stages of the task (encoding, retention, retrieval) and one for each of the three levels of difficulty (recalling of 2, 4, or 6 digits), convolved with a gamma hemodynamic response function. Additionally, we generated regressors of no interest for: trials in which participants answered incorrectly, rigid motion parameters and the top 10 aCompcor regressors obtained during preprocessing. First-level contrast parameter estimates (Copes) were used for higher level analyses to investigate fMRI activation at the group-level during Wmem processes. Only the retention phase of the WMem paradigm was analyzed, since it is difficult to dissociate attention from motion initiation processes secondary to encoding and retrieval phases, respectively. Single group average fMRI activation was assessed through a mixed effects (FLAME 1) model. Correction for multiple comparisons was carried out through random field theory (cluster forming threshold: *z >* 3.1), and regions were deemed significant if *p*_*cluster*_ *<* 0.05.

### 2.10. Association between Bold activity during retention phase and cognitive abilities

From identified clusters involved in the retention phase, we generated spherical regions of interest (ROI; 4mm radius) centered at the local maxima (Table 1). Considering previous reports of TLE patients displaying aberrant hippocampal activity during working memory (Campo et al., 2013; Stretton et al., 2013; Winston et al., 2013), we also included two spherical hippocampal ROIs derived from the DMN-TPN networks resulting from RS analyses. Mean BOLD percentage change during the retention phase relative to basal activity was obtained per ROI and was analyzed and compared between groups through Student’s t or Mann-Whitney U tests. Additionally, we examined whether such BOLD activity was correlated with neuropsychological performance by fitting ANCOVA models as described above for FC analysis. Each cognitive score was predicted as a function of BOLD activity per ROI and group membership. Similarly, we looked for group-specific correlations by fitting one ANCOVA model per ROI, with statistical significance considered at *p*_*F* *DR*_ *<* 0.05.

Finally, we explored whether regions involved in the retention phase of Sternberg’s memory task were functionally altered during rest. For this purpose each ROI was used to generate a whole-brain FC map through computation of Pearson’s correlation coefficient of the ROI’s time series and the time series of every other voxel. FMRIB-FSL Randomize (version v.2.9) was used with 5000 permutations to determine *p* value followed by threshold-free cluster enhancement to detect significant clusters.

### 2.11. Data availability

All data, including raw MRI images (T1-weighted, resting-state fMRI and task-based fMRI), and cognitive scores, are freely-available at Openneuro (data set ds004469).^2^

## 3. Results

### 3.1. Cognitive abilities

Analysis of cognitive indices derived from WAIS-IV and WMS-IV batteries showed that control subjects obtained average scores according to the population norm. In contrast, scores of TLE patients reflected mild to moderate cognitive impairment, with significantly lower scores in all the cognitive domains as compared with healthy subjects (Figure 2). After Bonferroni correction, all differences remained significant with exception of PR and VWMI. Scores of TLE patients were lowest in indexes related to memory recall and/or retention of information.

**Figure 2:**
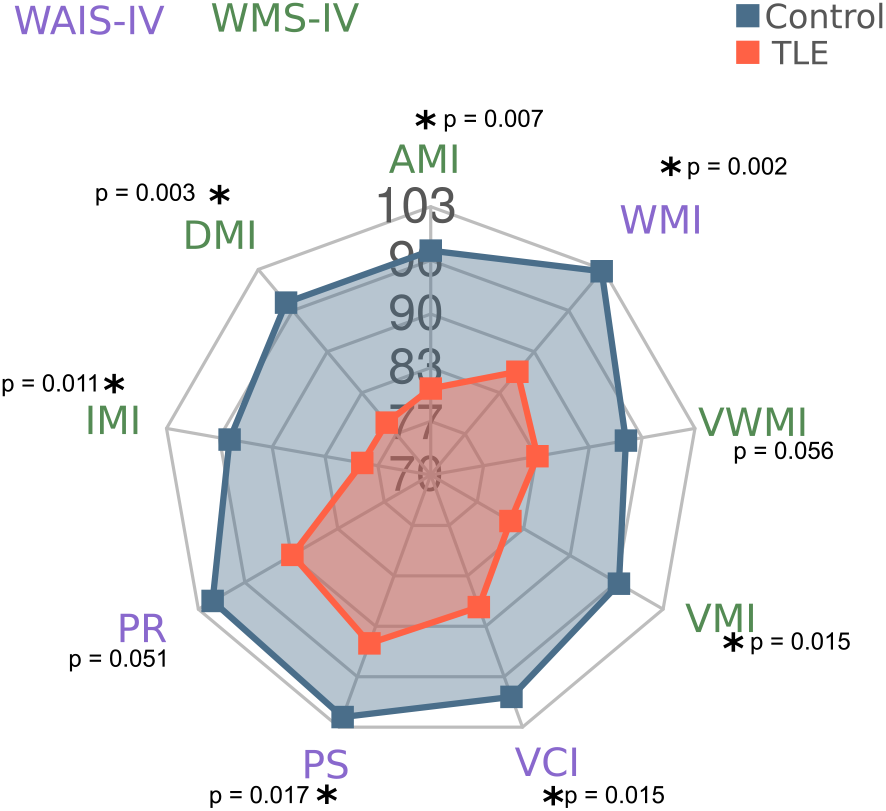
Cognitive indexes. Results are represented by group means. Significantly different indexes between groups are labeled with an asterisk (*) for corrected *p <* 0.05.

### 3.2. Network analysis

Analysis carried out with NBS revealed the presence of three components (sub-networks) that were significantly different between healthy participants and TLE patients. As shown in Figure 3A,B, Component 1 corresponds to a sub-network of the DMN which included both hippocampi, both middle temporal gyri, ventromedial prefrontal cortex and posterior cingulate cortex. Compared to healthy participants, TLE patients displayed a decrease of FC in this component. Similarly, TLE patients also showed a decrease in FC in Component 2 (Figure 3A,C), a sub-network of TPN, that involves nodes from the salience network (anterior insular cortices and paracingulate gyrus) and the contralateral supramarginal gyrus. On the other hand, while the control group showed the typical anticorrelation between DMN and TPN, patients with TLE did not show such divergent activity amongst these two networks (Figure 3A,D), involving structures also seen in Component 1, such as the ipsilateral hippocampus and middle temporal gyrus. In addition to the NBS approach, we searched for alterations in FC at individual connections through t-tests between groups (Figure 3E-G). After FDR correction, C.HIPP-I.HIPP and PCG-I.INS were significantly lower in TLE patients (*p*_*corr*_ *<* 5 *×* 10^*−*6^ for both).

**Figure 3:**
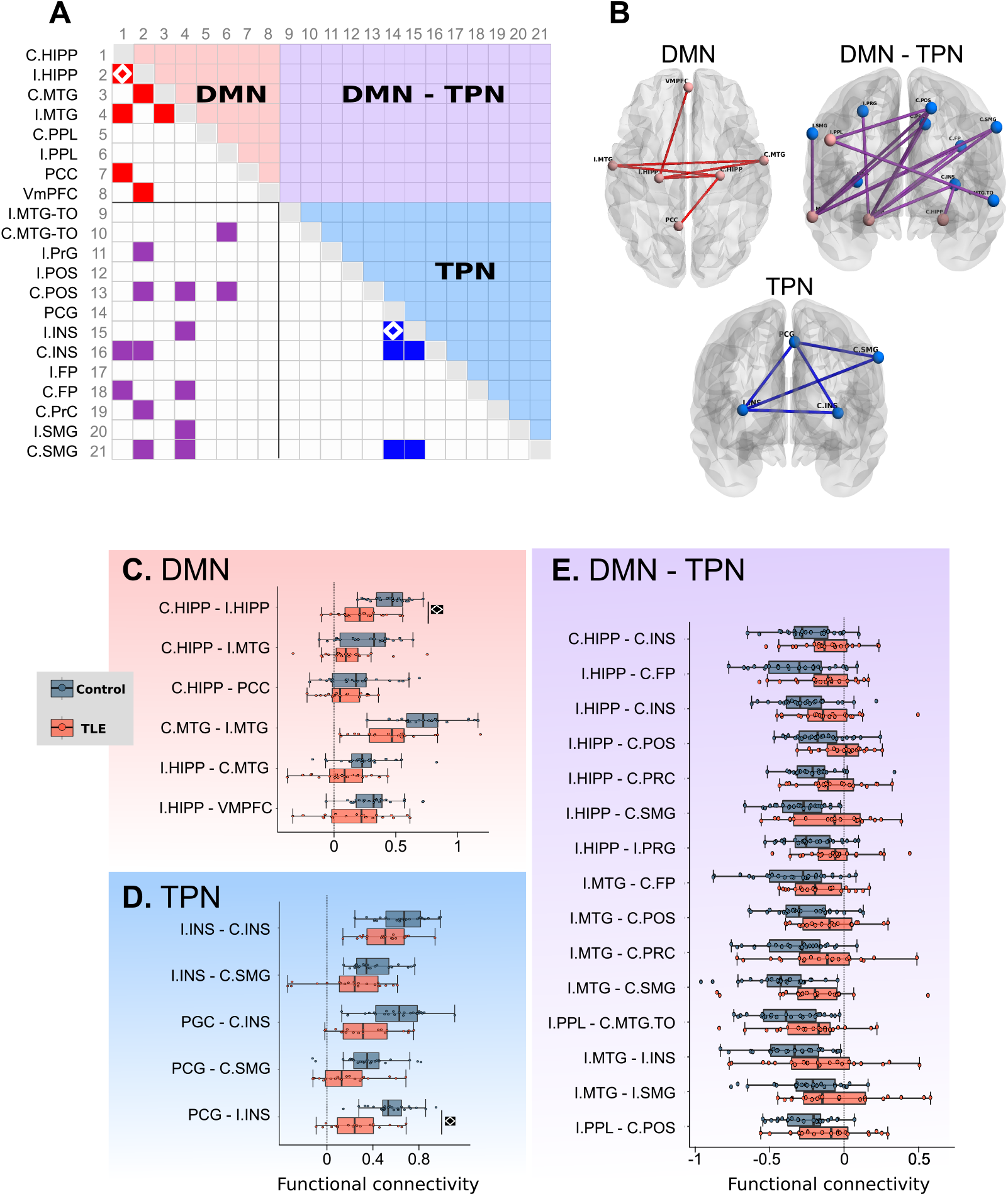
Functional connectivity differences between healthy subjects and TLE patients. A: Adjacency matrix showing between-group pairwise connectivity differences (filled cells), color-coded to identify three components identified with NBS analysis (default-mode network: DMN; task-positive networks: TPN; and their interaction: DMN-TPN. Their corresponding spatial configuration is shown in B. C-E: Boxplots show group-wise values at the edge level. Diamonds in A, and C-E represent significant between-group differences in pairwise node-node comparisons (FDR *p*_*corr*_ *<* 0.05). Refer to Table 1 for ROI abbreviations.

### 3.3. Associations between functional connectivity and cognitive abilities

Figure 4 shows significant group-specific associations between FC edges and cognitive abilities after correction for multiple comparisons. TLE patients showed several significant correlations between FC and cognitive abilities, while healthy subjects did not. These associations involved both hicompampi, middle temporal lobe regions, insular cortices and Ipsilateral precentral gyrus

**Figure 4:**
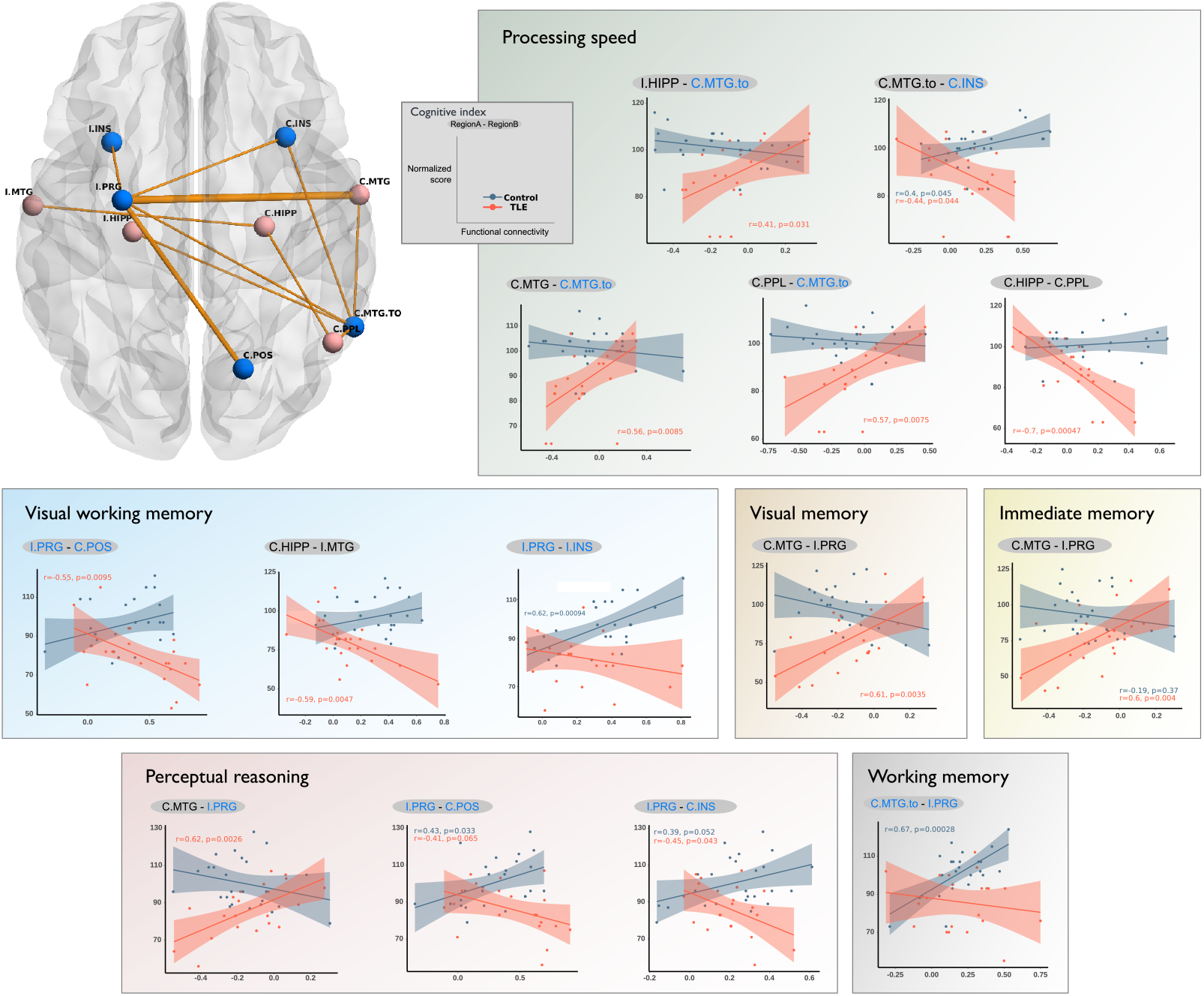
Significant group interactions between FC and cognitive abilities. Spatial representation of edges associated with cognition: Light red spheres represent nodes from the DMN, while blue spheres represent nodes from the TPN. Thickness of sticks is proportional to the number of associations between that particular edge with the cognitive indexes. Scatterplots are shown for statistically significant group interactions, grouped by cognitive index. The gray inset indicates the design for all plots; functional connectivity expressed as Fisher’s *z*. Pearson coefficients and *p* values are shown color-coded for significant group-specific correlations.

### 3.4. Sternberg’s memory task

#### 3.4.1. Behavioral paradigm

Control group and TLE patients did not differ in the number of correct responses (F(1,67) = 2.23, *p* = 0.139). Nonetheless, a significant effect of cognitive load (number of digits retained in memory) was found (F(2,134) = 7.991, *p <* 0.001). Post-hoc Bonferroni-corrected tests revealed that both groups performed significantly better when they had to memorize 2 vs 6 digits (t(32) = 2.97, *p <* 0.05 ; t(35) = 3.01, *p <* 0.05). In addition, the control group also performed significantly better when they had to memorize 2 vs 4 digits (t(32) = 2.97, *p <* 0.05). No interaction between group×digits was found (F(2,134) = 0.744, *p* = 0.47). We analyzed the mean reaction time of subjects along correct response trials. Mixed-effects ANOVA revealed significant effects of group (F(1,47) = 6.44, *p <* 0.05) and cognitive load (F(2,94) = 17.102, *p <* 0.0001) but no significant interaction between these factors (F(2,94) = 2.942, *p* = 0.058). Post-hoc comparisons revealed that in comparison to controls, TLE patients had significantly greater reaction times when they had to memorize either 2 digits (t(47) = -3.39, *p <* 0.01) or 4 digits (t(47) = -2.09, *p <* 0.05). Additionally, post-hoc tests revealed that only control subjects significantly increased their reaction time in the 4 (t(20) = -4.37, *p <* 0.001) and 6 digits (t(20) = -5.77, *p <* 0001) conditions in comparison with the 2 digits condition. Figure 5 illustrates these differences.

**Figure 5:**
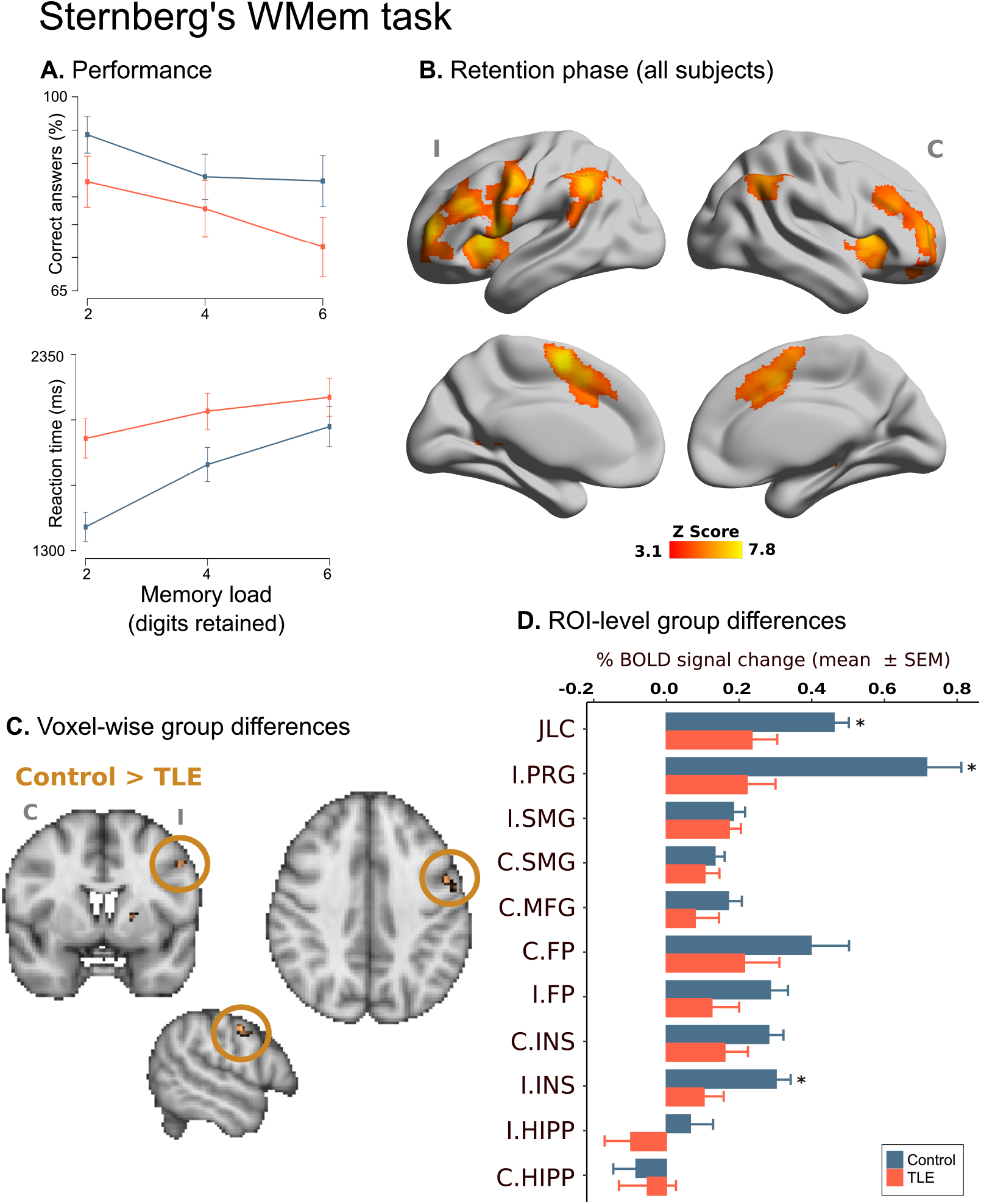
Sternberg’s working memory task. A: Correct answers and response time (mean and standard error) as a function of memory load. B: Group-level fMRI activation (controls and patients) during retention phase. Statistical map is shown referenced to the seizure onset side (I: ipsilateral; C: contralateral) of the epileptogenic focus. C: Significant between-group voxelwise differences on fMRI activation during retention phase. D: Between-group differences in mean percentage of BOLD signal change in ROIs derived from retention phase activation map. Asterisks denote *p*_*F* *DR*_ *<* 0.05.

#### 3.4.2. Functional correlates of Sternberg memory task

We fitted a GLM to find the significant brain regions involved in the retention phase of the Wmem task. In both groups, we observed bilateral activation of frontal poles, supramarginal gyri, anterior insular cortices and middle frontal gyri (Figure 5). Furthermore we detected activation in the Ipsilateral (left) precentral gyrus, and the Juxtapositional Lobule cortex/Paracingulate gyrus. When comparing between-group activity, planned contrasts revealed that TLE patients had less BOLD activity than controls in ipsilateral precentral gyrus. Contrarily, contralateral frontal operculum had significantly greater activity in TLE patients. In all cases, controls display greater BOLD signal change as compared to TLE patients.

#### 3.4.3. Association between BOLD activity during retention phase and cognitive abilities

Voxelwise analyses revealed that correlation between cognitive indexes and BOLD activity within supramarginal gyri, intraparietal sulci, ipsilateral precentral gyrus and contralateral occipital cortex significantly differed between groups. Figure 6 illustrates correlations between mean bold percentage change during Sternberg’s task and cognitive indexes that differed between patients and controls. Group interaction showed different directions of slopes, in all cases showing patients having positive correlations between the two variables.

**Figure 6:**
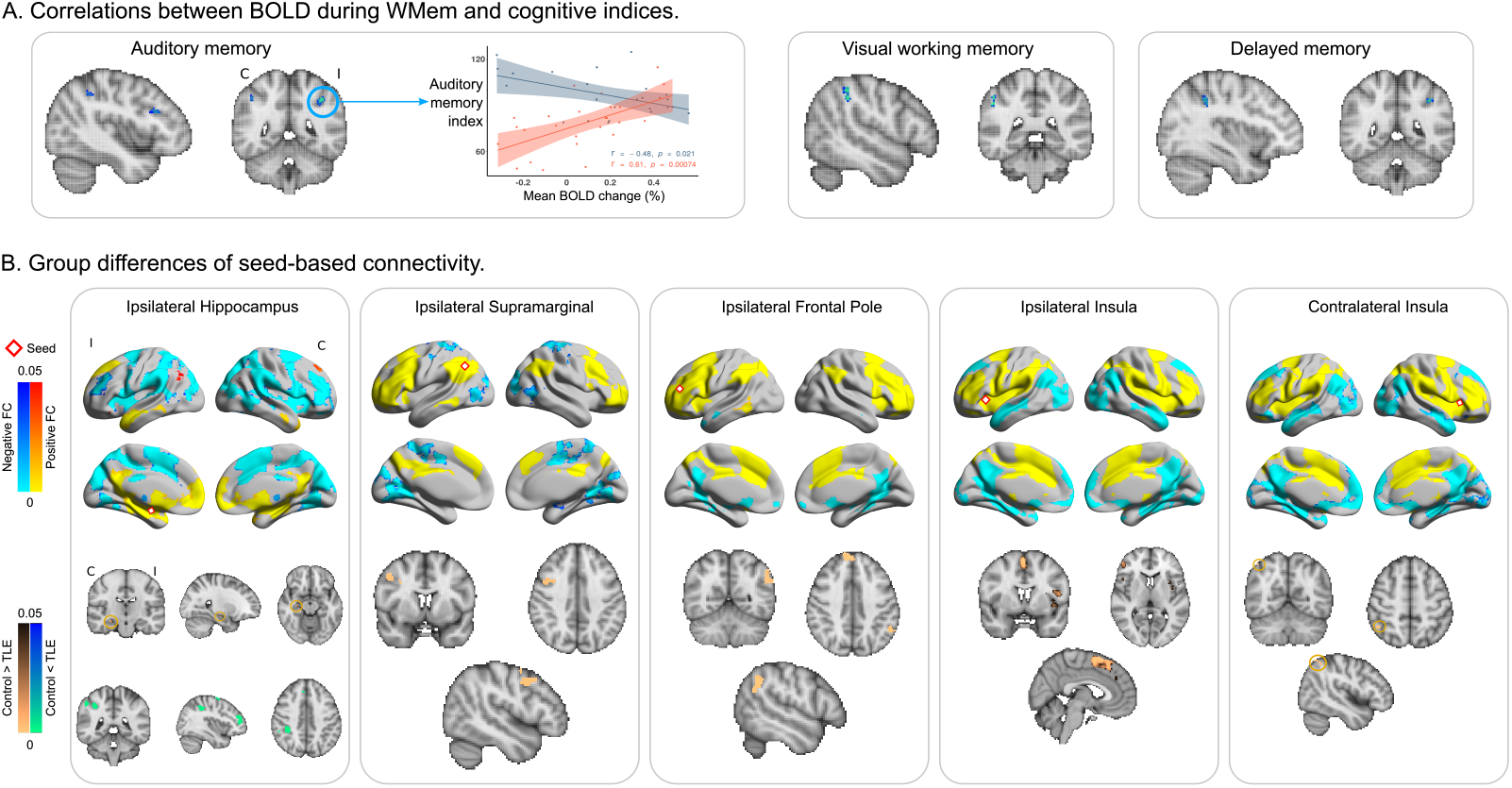
A: Correlations between BOLD activity and cognitive scores modulated by Group. ANCOVAs showed positive correlations only in TLE patients, and not in controls for three different memory indices. Analysis was restricted to regions involved in Sternberg’s task, and shown corrected for family-wise errors. A scatter plot is shown for illustrative purposes for auditory memory index. B: Voxelwise analyses of seed-based RSFC for ROIs derived from the retention stage of Sternberg’s memory task. Top row shows connectivity of the whole brain RSFC of seed (red-outline diamonds). Bottom row shows group-wise differences of the corresponding connectivity maps.

#### 3.4.4. Alterations in RS-FC in retention phase (ROI-analysis)

Finally, we hypothesized that TLE patients would display alterations of RSFC related to regions involved in the retention phase of Sternberg’s memory task. To this end, we used seed-based connectivity analyses (SCA) and evaluated the correlation of the BOLD signal between the seed region and the rest of the brain. Brain areas found to be active in the retention phase of the Wmem task (Figure 6) and both hippocampi served as seed regions. We found significant group-level differences in the connectivity of the following areas: both insular cortices, both hippocampi, ipsilateral frontal pole and ipsilateral supramarginal gyrus. Roughly, we observed that patients had decreased RSFC in salience network (as evidenced by insular SCA) and ipsilateral fronto-parietal network (as evidenced by ipsilateral supramarginal and ipsilateral frontal pole SCA). Large between-group differences were found in hippocampal SCA. In general, patients displayed decreased RSFC between C.HIPP and I.HIPP and greater RSFC between C.HIPP and Contralateral fronto-parietal network regions. All of these results were consistent and similar to those obtained through the network-based approach, showing abnormal RSFC in TPN, DMN and in anticorrelations between both systems.

## 4. Discussion

Cognitive deficits are common in patients with TLE and affect several cognitive domains that involve brain regions within and beyond the temporal lobes. Here, we confirm that, as a group, TLE patients have cognitive deficits across domains, albeit with considerable inter-subject heterogeneity. Through analysis of resting-state fMRI we showed that cognitive deficits are associated with disorganized functional connectivity, with patients showing decreased connectivity within the default-mode and temporo-polar networks and, contrastingly, a less marked decoupling between DMN and TPN than what is typically seen in healthy individuals. Extensive cognitive testing allowed us to identify that the degree of specific region-to-region connectivity is associated with neuropsychometric scores of visual memory, processing speed, and perceptual reasoning. Further testing of working memory within the imaging session showed that alterations of WMem are accompanied by reduced activity of temporo-parietal regions, which are also less tightly connected to remote brain areas.

Cognitive performance is not homogeneous across TLE patients, who show cognitive abilities ranging from intact to severe generalized cognitive deficits (Hermann et al., 2007; Rodríguez-Cruces et al., 2020). The range and type of cognitive disabilities seen in TLE patients have in turn been associated with specific patterns of morphological alterations (Dabbs et al., 2009) and structural connectivity (Hermann et al., 2021; Rodríguez-Cruces et al., 2020). Therefore, it is believed that the distributed gray (Whelan et al., 2018) and white matter damage (Hatton et al., 2020) seen in TLE explain the heterogeneity of the type and severity of cognitive deficits in these patients through a disruption of the normal flow of information within the brain. Indeed, abnormalities of functional connectivity have been reported in TLE, providing evidence for abnormalities of network topography (Liao et al., 2010a,b) and alterations of the DMN, the attention network, and the reward/emotion network (Bernhardt et al., 2016; Larivi`ere et al., 2020; Morgan et al., 2015; Qin et al., 2020; Vlooswijk et al., 2011; Voets et al., 2009; Zhang et al., 2009). Spatial patterns of rsfMRI abnormalities greatly coincide with structural damage (Caciagli et al., 2014; Voets et al., 2012), and appear to be mediated by microstructural abnormalities of white matter (Larivi`ere et al., 2020). Normally, TPN increase their activity during cognitive tasks, and are negatively correlated with the DMN (Fox et al., 2005). Our results recapitulate the previously observed reduction of functional connectivity within each of the DMN and TPN. Moreover, TLE patients failed to show anticorrelations between DMN and TPN, while healthy participants displayed said typical association (Figure 3) (Fornito et al., 2012; Konishi et al., 2015). In agreement with previous reports, inter-hippocampal connectivity was decreased in TLE patients, as was the interaction between the ipsilateral hippocampus and temporal neocortex (Doucet et al., 2013; Maccotta et al., 2013; Morgan et al., 2011).

We found several instances where TLE and healthy participants displayed different associations between region-to-region connectivity and cognitive performance (Figure 4). The majority of these between-group differences consisted of significant correlations seen in patients, but not in controls. It is tempting to associate increased functional connectivity with better performance, and there are examples advocating for that view in our results, and in the literature (Wagner et al., 2007). Nonetheless, the opposite can be observed in some connectivity edges. In our study we found that visual working memory was poorer in TLE patients with higher degree of connectivity between the contralateral hippocampus and ipsilateral temporal neocortex, and between the ipsilateral motor and parieto-occipital cortices. Similarly, increased connectivity between ipsilateral insula and motor cortex had a negative impact on visual working memory and perceptual reasoning, while connectivity between the contralateral insula and temporal neocortex affected processing speed. Perceptual reasoning was negatively impacted by increased connectivity within the TPN, but improved by greater connectivity between contralateral temporal neocortex and ipsilateral motor cortex. These associations not seen in controls suggest compensatory mechanisms at play in patients to maintain cognitive abilities (Voets et al., 2012). A similar reconfiguration of brain networks related to WMem has also been reported in infants with periventricular leukomalacia (García-Gomar et al., 2013).

There are discrepancies in our results as compared to previous reports. First, we do not see an increase of connectivity of the contralateral mesiotemporal networks, which has been shown to correlate with WMem performance (Bettus et al., 2009). Increased functional connectivity between ipsilateral mesial temporal structures and the ipsilateral posterior DMN have been reported to have an association with poorer verbal memory abilities (Doucet et al., 2013; Holmes et al., 2014), while increased coupling between the ipsilateral hippocampus and contralateral posterior DMN shows a positive relation with improved verbal memory (Holmes et al., 2014). In our study, however, the only association between the connectivity of ipsilateral hippocampus and cognitive abilities was related to the contralateral MTG and improved processing speed. Also, the contralateral left mesial temporal lobe and the ipsilateral mesial prefrontal cortex was positively associated with higher scores for non-verbal memory (Doucet et al., 2013). In our cohort, contralateral hippocampus connectivity to ipsilateral MTG and contralateral PPL correlated negatively with VWMI and processing speed, respectively. While some of the previous studies studied left and right TLE patients separately, we pooled both patient groups and studied their brains according to ipsi- or contralateral to seizure onset. This, added to the known functional brain asymmetries behind many cognitive abilities (e.g., verbal) likely account for the differences between studies.

Despite having clear deficiencies in working memory when evaluated outside the scanner (Figure 2), TLE patients in our study showed only a tendency for reduced correct answers during Sternberg’s task in the MRI scanner, but there was a clear difference between the two groups in their reaction times, with patients responding much later than controls, even in trials with minimal cognitive load (Figure 5). Correspondingly, the large majority of brain regions that form the normal fronto-parietal network involved in WMem (Owen et al., 2005) showed less activation in TLE patients than healthy participants, replicating and extending previous findings (Stretton et al., 2012). Moreover, the level of activation of fronto-parietal regions involved in WMem showed differential correlations with other immediate and delayed memory indices, with positive correlations seen in TLE patients, but not in healthy participants (Figure 6), suggesting compensatory mechanisms to maintain WMem. Finally, seed-based analyses of resting-state BOLD activity showed larger connectivity between the hippocampi and fronto-parietal regions in TLE patients, as compared to controls (Figure 6). This result is in line with the increased co-activation of these same regions seen during an N-back WMem task (Stretton et al., 2013). In said report, as in ours, increased co-activation of these brain regions was associated with poor WMem performance. Our work extends said observations by showing that TLE patients have decreased connectivity between ipsilateral insula and frontal regions, and between ipsilateral frontal pole and supramarginal gyrus. Thus, our results show the WMem network of TLE patients is altered at rest, is suboptimally engaged during memory retention, and is differentially modulated to subserve immediate and delayed memory performance. There are some limitations in our study. First, we could not obtain histological confirmation of hippocampal sclerosis, as the majority of patients studied, unfortunately, did not undergo surgical treatment. Despite best efforts from the medical community, there are reduced opportunities for surgical treatment of TLE in low- and middle-income countries (Watila et al., 2019). Also, in an effort to boost statistical power, we congregated TLE patients regardless of the hemisphere of seizure onset, and analyzed their data according to ipsilateral or contralateral to the epileptogenic temporal lobe. This strategy makes it difficult to adequately interpret results related to cognitive or functional hemispheric specialization, such as verbal skills in our resting-state connectivity analyses. On the other hand, findings regarding the bilateral and symmetrical network involved in WMem are likely less affected by this limitation.

In conclusion, our results allowed us to link whole-brain connectivity with cognitive abilities in multiple domains, showing abnormal patterns in TLE patients that tie together with cognitive deficits, as well as putative compensatory mechanisms. Targeting one specific cognitive feature, namely WMem, we were able to identify abnormalities in the activity of the underlying brain regions in TLE, but also how these associate to other brain regions and networks and affect several other cognitive abilities.

## Data Availability

All data, including raw MRI images (T1-weighted, resting-state fMRI and task-based fMRI), and cognitive scores, are freely-available at Openneuro (data set ds004469; https://doi.org/10.18112/openneuro.ds004469.v1.0.0)

## Acknowledgements

We thank the patients and control subjects for their willingness to participate, the medical specialists who helped us with their recruitment, and the clinical personnel at the National Laboratory for magnetic resonance imaging. We are grateful to Juan Ortíz-Retana and Leopoldo González-Santos for technical assistance. We thank the National Laboratory for scientific visualization (LAVIS) and staff, namely Luis Aguilar and Alejandro de León. We are grateful to the many people who have at some point participated in this project: Leticia Velázquez-Pérez, David Trejo, Héctor Barragán, Arturo Domínguez, Ildefonso Rodríguez-Leyva, Ana Luisa Velasco, Luis Octavio Jiménez, Daniel Atilano, Elizabeth González Olvera, Rafael Moreno, and Ana Elena Rosas.

## Footnotes

2 https://doi.org/10.18112/openneuro.ds004469.v1.0.0

## References

Agah, E., Asgari-Rad, N., Ahmadi, M., Tafakhori, A., & Aghamollaii, V. (2017). Evaluating executive function in patients with temporal lobe epilepsy using the frontal assessment battery. Epilepsy Research, 133, 22–27. doi:10.1016/j.eplepsyres.2017.03.011.

Andrews-Hanna, J. R., Reidler, J. S., Sepulcre, J., Poulin, R., & Buckner, R. L. (2010). Functional-anatomic fractionation of the brain’s default network. Neuron, 65, 550–562. doi:10.1016/j.neuron.2010.02.005.

Baddeley, A. (2012). Working memory: Theories, models, and controversies. Annual Review of Psychology, 63, 1–29. doi:10.1146/annurev-psych-120710-100422.

Bernhardt, B. C., Bernasconi, A., Liu, M., Hong, S.-J., Caldairou, B., Goubran, M., Guiot, M. C., Hall, J., & Bernasconi, N. (2016). The spectrum of structural and functional imaging abnormalities in temporal lobe epilepsy. Annals of Neurology, 80, 142–153. doi:10.1002/ana.24691.

Bettus, G., Guedj, E., Joyeux, F., Confort-Gouny, S., Soulier, E., Laguitton, V., Cozzone, P. J., Chauvel, P., Ranjeva, J.-P., Bartolomei, F., & et al. (2009). Decreased basal fmri functional connectivity in epileptogenic networks and contralateral compensatory mechanisms. Human Brain Mapping, 30, 1580–1591. doi:10.1002/hbm.20625.

Black, L. C., Schefft, B. K., Howe, S. R., Szaflarski, J. P., Yeh, H.-s., & Privitera, M. D. (2010). The effect of seizures on working memory and executive functioning performance. Epilepsy & Behavior, 17, 412–419. doi:10.1016/j.yebeh.2010.01.006.

Caciagli, L., Bernhardt, B. C., Hong, S.-J., Bernasconi, A., & Bernasconi, N. (2014). Functional network alterations and their structural substrate in drug-resistant epilepsy. Frontiers in Neuroscience, 8. URL: https://www.frontiersin.org/article/10.3389/fnins.2014.00411.

Campo, P., Garrido, M. I., Moran, R. J., García-Morales, I., Poch, C., Toledano, R., Gil-Nagel, A., Dolan, R. J., & Friston, K. J. (2013). Network reconfiguration and working memory impairment in mesial temporal lobe epilepsy. NeuroImage, 72, 48–54. doi:10.1016/j.neuroimage.2013.01.036.

Craddock, C., Sikka, S., Brian, B., Khanuja, R., Ghosh, S., Yan, C., Li, Q., Lurie, D., Vogelstein, J., Burns, R., & et al. (2013). Towards automated analysis of connectomes: The configurable pipeline for the analysis of connectomes (c-pac). Frontiers in Neuroinformatics, 7. URL: http://www.frontiersin.org/10.3389/conf.fninf.2013.09.00042/event_abstract. doi:10.3389/conf.fninf.2013.09.00042.

Dabbs, K., Jones, J., Seidenberg, M., & Hermann, B. (2009). Neuroanatomical correlates of cognitive phenotypes in temporal lobe epilepsy. Epilepsy & Behavior, 15, 445–451. doi:10.1016/j.yebeh.2009.05.012.

Delaney, R. C., Rosen, A. J., Mattson, R. H., & Novelly, R. A. (1980). Memory function in focal epilepsy: A comparison of non-surgical, unilateral temporal lobe and frontal lobe samples. Cortex, 16, 103–117. doi:10.1016/S0010-9452(80)80026-8.

Deleo, F., Thom, M., Concha, L., Bernasconi, A., Bernhardt, B. C., & Bernasconi, N. (2018). Histological and mri markers of white matter damage in focal epilepsy. Epilepsy Research, 140, 29–38. doi:10.1016/j.eplepsyres.2017.11.010.

Doucet, G., Osipowicz, K., Sharan, A., Sperling, M. R., & Tracy, J. I. (2013). Extratemporal functional connectivity impairments at rest are related to memory performance in mesial temporal epilepsy. Human Brain Mapping, 34, 2202–2216. doi:10.1002/hbm.22059.

Eddy, C. M., Rickards, H. E., & Cavanna, A. E. (2011). The cognitive impact of antiepileptic drugs. Therapeutic Advances in Neurological Disorders, 4, 385–407. doi:10.1177/1756285611417920.

Eriksson, J., Vogel, E. K., Lansner, A., Bergström, F., & Nyberg, L. (2015). Neurocognitive architecture of working memory. Neuron, 88, 33–46. doi:10.1016/j.neuron.2015.09.020.

Esteban, O., Markiewicz, C. J., Blair, R. W., Moodie, C. A., Isik, A. I., Erramuzpe, A., Kent, J. D., Goncalves, M., DuPre, E., Snyder, M., & et al. (2019). fmriprep: a robust preprocessing pipeline for functional mri. Nature Methods, 16, 111–116. doi:10.1038/s41592-018-0235-4.

Fisher, R. S., Vickrey, B. G., Gibson, P., Hermann, B., Penovich, P., Scherer, A., & Walker, S. (2000). The impact of epilepsy from the patient’s perspective i. descriptions and subjective perceptions. Epilepsy Research, 41, 39–51. doi:10.1016/S0920-1211(00)00126-1.

Fornito, A., Harrison, B. J., Zalesky, A., & Simons, J. S. (2012). Competitive and cooperative dynamics of large-scale brain functional networks supporting recollection. Proceedings of the National Academy of Sciences, 109, 12788–12793. doi:10.1073/pnas.1204185109.

Fox, M. D., Snyder, A. Z., Vincent, J. L., Corbetta, M., Van Essen, D. C., & Raichle, M. E. (2005). The human brain is intrinsically organized into dynamic, anticorrelated functional networks. Proceedings of the National Academy of Sciences, 102, 9673–9678. doi:10.1073/pnas.0504136102.

Friston, K. J., Williams, S., Howard, R., Frackowiak, R. S. J., & Turner, R. (1996). Movement-related effects in fmri time-series. Magnetic Resonance in Medicine, 35, 346–355. doi:10.1002/mrm.1910350312.

García-Gomar, M. L., Santiago-Rodríguez, E., Rodríguez-Camacho, M., & Harmony, T. (2013). Visuospatial working memory in toddlers with a history of periventricular leukomalacia: An eeg narrow-band power analysis. PLoS ONE, 8, e69837. doi:10.1371/journal.pone.0069837.

Giovagnoli, A. R., & Avanzini, G. (2000). Quality of life and memory performance in patients with temporal lobe epilepsy. Acta Neurologica Scandinavica, 101, 295–300. doi:10.1034/j.1600-0404.2000.90257a.x.

Hatton, S. N., Huynh, K. H., Bonilha, L., Abela, E., Alhusaini, S., Altmann, A., Alvim, M. K. M., Balachandra, A. R., Bartolini, E., Bender, B., & et al. (2020). White matter abnormalities across different epilepsy syndromes in adults: an enigma-epilepsy study. Brain: A Journal of Neurology, 143, 2454–2473. doi:10.1093/brain/awaa200.

Hermann, B., Seidenberg, M., Lee, E.-J., Chan, F., & Rutecki, P. (2007). Cognitive phenotypes in temporal lobe epilepsy. Journal of the International Neuropsychological Society, 13, 12–20. doi:10.1017/S135561770707004X.

Hermann, B. P., Struck, A. F., Busch, R. M., Reyes, A., Kaestner, E., & McDonald, C. R. (2021). Neurobehavioural comorbidities of epilepsy: towards a network-based precision taxonomy. Nature Reviews Neurology, 17, 731–746. doi:10.1038/s41582-021-00555-z.

Holmes, M., Folley, B. S., Sonmezturk, H. H., Gore, J. C., Kang, H., Abou-Khalil, B., & Morgan, V. L. (2014). Resting state functional connectivity of the hippocampus associated with neurocognitive function in left temporal lobe epilepsy. Human Brain Mapping, 35, 735–744. doi:10.1002/hbm.22210.

Hoppe, C., Elger, C. E., & Helmstaedter, C. (2007). Long-term memory impairment in patients with focal epilepsy. Epilepsia, 48, 26–29.

Hudson, J. M., Flowers, K. A., & Walster, K. L. (2014). Attentional control in patients with temporal lobe epilepsy. Journal of Neuropsychology, 8, 140–146. doi:10.1111/jnp.12008.

Khalife, M. R., Scott, R. C., & Hernan, A. E. (2022). Mechanisms for cognitive impairment in epilepsy: Moving beyond seizures. Frontiers in Neurology, 13. URL: https://www.frontiersin. org/article/10.3389/fneur.2022.878991.

Konishi, M., McLaren, D. G., Engen, H., & Smallwood, J. (2015). Shaped by the past: The default mode network supports cognition that is independent of immediate perceptual input. PLOS ONE, 10, e0132209. doi:10.1371/journal.pone.0132209.

Larivìere, S., Weng, Y., Vos de Wael, R., Royer, J., Frauscher, B., Wang, Z., Bernasconi, A., Bernasconi, N., Schrader, D. V., Zhang, Z., & et al. (2020). Functional connectome contractions in temporal lobe epilepsy: Microstructural underpinnings and predictors of surgical outcome. Epilepsia, 61, 1221–1233. doi:10.1111/epi.16540.

Liao, W., Zhang, Z., Pan, Z., Mantini, D., Ding, J., Duan, X., Luo, C., Lu, G., & Chen, H. (2010a). Altered functional connectivity and small-world in mesial temporal lobe epilepsy. PLOS ONE, 5, e8525. doi:10.1371/journal.pone.0008525.

Liao, W., Zhang, Z., Pan, Z., Mantini, D., Ding, J., Duan, X., Luo, C., Wang, Z., Tan, Q., Lu, G., & et al. (2010b). Default mode network abnormalities in mesial temporal lobe epilepsy: A study combining fmri and dti. Human Brain Mapping,. URL: http://www.ncbi.nlm.nih.gov.login.ezproxy.library.ualberta.ca/pubmed/20533558. doi:10.1002/hbm.21076.

Maccotta, L., He, B. J., Snyder, A. Z., Eisenman, L. N., Benzinger, T. L., Ances, B. M., Corbetta, M., & Hogan, R. E. (2013). Impaired and facilitated functional networks in temporal lobe epilepsy. NeuroImage: Clinical, 2, 862–872. doi:10.1016/j.nicl.2013.06.011.

Mansouri, F. A., Rosa, M. G. P., & Atapour, N. (2015). Working memory in the service of executive control functions. Frontiers in Systems Neuroscience, 9, 166. doi:10.3389/fnsys.2015.00166.

Milner, B. (1972). Disorders of learning and memory after temporal lobe lesions in man. Clin Neurosurg, 19, 421—446.

Morgan, V. L., Abou-Khalil, B., & Rogers, B. P. (2015). Evolution of functional connectivity of brain networks and their dynamic interaction in temporal lobe epilepsy. Brain Connectivity, 5, 35–44. doi:10.1089/brain.2014.0251.

Morgan, V. L., Rogers, B. P., Sonmezturk, H. H., Gore, J. C., & Abou-Khalil, B. (2011). Cross hippocampal influence in mesial temporal lobe epilepsy measured with high temporal resolution functional magnetic resonance imaging. Epilepsia, 52, 1741–1749. doi:10.1111/j.1528-1167.2011.03196.x.

Owen, A. M., McMillan, K. M., Laird, A. R., & Bullmore, E. (2005). N-back working memory paradigm: a meta-analysis of normative functional neuroimaging studies. Human Brain Mapping, 25, 46–59. doi:10.1002/hbm.20131.

Power, J. D., Barnes, K. A., Snyder, A. Z., Schlaggar, B. L., & Petersen, S. E. (2012). Spurious but systematic correlations in functional connectivity mri networks arise from subject motion. NeuroImage, 59, 2142–2154. doi:10.1016/j.neuroimage.2011.10.018.

Qin, L., Jiang, W., Zheng, J., Zhou, X., Zhang, Z., & Liu, J. (2020). Alterations functional connectivity in temporal lobe epilepsy and their relationships with cognitive function: A longitudinal resting-state fmri study. Frontiers in Neurology, 11. URL: https://www.frontiersin.org/ article/10.3389/fneur.2020.00625.

Rodriguez-Cruces, R., Royer, J., Larivìere, S., Bassett, D. S., Caciagli, L., & Bernhardt, B. C. (2022). Multimodal connectome biomarkers of cognitive and affective dysfunction in the common epilepsies. Network Neuroscience, 6, 320–338. doi:10.1162/netn_a_00237.

Rodríguez-Cruces, R., Bernhardt, B. C., & Concha, L. (2020). Multidimensional associations between cognition and connectome organization in temporal lobe epilepsy. NeuroImage, 213, 116706. doi:10.1016/j.neuroimage.2020.116706.

Rodríguez-Cruces, R., Velázquez-Pérez, L., Rodríguez-Leyva, I., Velasco, A. L., Trejo-Martínez, D., Barragán-Campos, H. M., Camacho-Téllez, V., & Concha, L. (2018). Association of white matter diffusion characteristics and cognitive deficits in temporal lobe epilepsy. Epilepsy & Behavior, 79, 138–145. doi:10.1016/j.yebeh.2017.11.040.

Satterthwaite, T. D., Elliott, M. A., Gerraty, R. T., Ruparel, K., Loughead, J., Calkins, M. E., Eickhoff, S. B., Hakonarson, H., Gur, R. C., Gur, R. E., & et al. (2013). An improved framework for confound regression and filtering for control of motion artifact in the preprocessing of resting-state functional connectivity data. NeuroImage, 64, 240–256. doi:10.1016/j.neuroimage.2012.08.052.

Scoville, W. B., & Milner, B. (1957). Loss of recent memory after bilateral hippocampal lesions. J Neurol Neurosurg Psychiatry, 20, 11—21.

Smallwood, J., Brown, K., Baird, B., & Schooler, J. W. (2012). Cooperation between the default mode network and the frontal–parietal network in the production of an internal train of thought. Brain Research, 1428, 60–70. doi:10.1016/j.brainres.2011.03.072.

Smith, S. M., Jenkinson, M., Woolrich, M. W., Beckmann, C. F., Behrens, T. E. J., Johansen-Berg, H., Bannister, P. R., De Luca, M., Drobnjak, I., Flitney, D. E., & et al. (2004). Advances in functional and structural mr image analysis and implementation as fsl. NeuroImage, 23 Suppl 1, S208–219. doi:10.1016/j.neuroimage.2004.07.051.

Stretton, J., Winston, G., Sidhu, M., Bonelli, S., Centeno, M., Vollmar, C., Cleary, R., Williams, E., Symms, M., Koepp, M., & et al. (2013). Disrupted segregation of working memory networks in temporal lobe epilepsy. NeuroImage: Clinical, 2, 273–281. doi:10.1016/j.nicl.2013.01.009.

Stretton, J., Winston, G., Sidhu, M., Centeno, M., Vollmar, C., Bonelli, S., Symms, M., Koepp, M., Duncan, J. S., & Thompson, P. J. (2012). Neural correlates of working memory in temporal lobe epilepsy — an fmri study. NeuroImage, 60, 1696–1703. doi:10.1016/j.neuroimage.2012.01.126.

Tudesco, I. d. S. S., Vaz, L. J., Mantoan, M. A. S., Belzunces, E., Noffs, M. H., Caboclo, L. O. S. F., Yacubian, E. M. T., Sakamoto, A. C., & Bueno, O. F. A. (2010). Assessment of working memory in patients with mesial temporal lobe epilepsy associated with unilateral hippocampal sclerosis. Epilepsy & Behavior, 18, 223–228. doi:10.1016/j.yebeh.2010.04.021.

Téllez-Zenteno, J. F., & Hernández-Ronquillo, L. (2012). A review of the epidemiology of temporal lobe epilepsy. Epilepsy research and treatment, 2012, 630853. doi:10.1155/2012/630853.

Vlooswijk, M. C. G., Jansen, J. F. A., Jeukens, C. R. L. P. N., Marian Majoie, H. J., Hofman, P. A. M., de Krom, M. C. T. F. M., Aldenkamp, A. P., & Backes, W. H. (2011). Memory processes and prefrontal network dysfunction in cryptogenic epilepsy. Epilepsia, 52, 1467–1475. doi:10.1111/j.1528-1167.2011.03108.x.

Voets, N. L., Adcock, J. E., Stacey, R., Hart, Y., Carpenter, K., Matthews, P. M., & Beckmann, C. F. (2009). Functional and structural changes in the memory network associated with left temporal lobe epilepsy. Human Brain Mapping, 30, 4070–4081. doi:10.1002/hbm.20830.

Voets, N. L., Beckmann, C. F., Cole, D. M., Hong, S., Bernasconi, A., & Bernasconi, N. (2012). Structural substrates for resting network disruption in temporal lobe epilepsy. Brain, 135, 2350–2357. doi:10.1093/brain/aws137.

Wagner, K., Frings, L., Halsband, U., Everts, R., Buller, A., Spreer, J., Zentner, J., & Schulze-Bonhage, A. (2007). Hippocampal functional connectivity reflects verbal episodic memory network integrity. NeuroReport, 18, 1719–1723. doi:10.1097/WNR.0b013e3282f0d3c5.

Watila, M. M., Xiao, F., Keezer, M. R., Miserocchi, A., Winkler, A. S., McEvoy, A. W., & Sander, J. W. (2019). Epilepsy surgery in low-and middle-income countries: A scoping review. Epilepsy & Behavior: E&B, 92, 311–326. doi:10.1016/j.yebeh.2019.01.001.

Whelan, C. D., Altmann, A., Botía, J. A., Jahanshad, N., Hibar, D. P., (…), Concha, L., (…), Rodriguez-Cruces, R. et al. (2018). Structural brain abnormalities in the common epilepsies assessed in a worldwide enigma study. Brain,. URL: https://academic.oup.com/brain/advance-article/doi/10.1093/brain/awx341/4818311. doi:10.1093/brain/awx341.

Winston, G. P., Stretton, J., Sidhu, M. K., Symms, M. R., Thompson, P. J., & Duncan, J. S. (2013). Structural correlates of impaired working memory in hippocampal sclerosis. Epilepsia, 54, 1143–1153. doi:10.1111/epi.12193.

Yarkoni, T., Poldrack, R. A., Nichols, T. E., Van Essen, D. C., & Wager, T. D. (2011). Large-scale automated synthesis of human functional neuroimaging data. Nature Methods, 8, 665–670. doi:10.1038/nmeth.1635.

Zalesky, A., Fornito, A., & Bullmore, E. T. (2010). Network-based statistic: Identifying differences in brain networks. NeuroImage, 53, 1197–1207. doi:10.1016/j.neuroimage.2010.06.041.

Zamarian, L., Trinka, E., Bonatti, E., Kuchukhidze, G., Bodner, T., Benke, T., Koppelstaetter, F., & Delazer, M. (2011). Executive functions in chronic mesial temporal lobe epilepsy. Epilepsy Research and Treatment, 2011, 596174. doi:10.1155/2011/596174.

Zeman, A., Kapur, N., & Jones-Gotman, M. (2012). Epilepsy and memory. Oxford University Press.

Zhang, Z., Lu, G., Zhong, Y., Tan, Q., Liao, W., Chen, Z., Shi, J., & Liu, Y. (2009). Impaired perceptual networks in temporal lobe epilepsy revealed by resting fmri. Journal of Neurology, 256, 1705–1713. doi:10.1007/s00415-009-5187-2.

Zhao, F., Kang, H., You, L., Rastogi, P., Venkatesh, D., & Chandra, M. (2014). Neuropsychological deficits in temporal lobe epilepsy: A comprehensive review. Annals of Indian Academy of Neurology, 17, 374–382. doi:10.4103/0972-2327.144003.

